# Levels and determinants of COVID-19 vaccine hesitancy among sub-Saharan African adolescents

**DOI:** 10.1101/2022.05.18.22275274

**Authors:** Dongqing Wang, Angela Chukwu, Mary Mwanyika-Sando, Sulemana Watara Abubakari, Nega Assefa, Isabel Madzorera, Elena C Hemler, Abbas Ismail, Bruno Lankoande, Frank Mapendo, Ourohiré Millogo, Firehiwot Workneh, Temesgen Azemraw, Lawrence G Febir, Christabel James, Amani Tinkasimile, Kwaku Poku Asante, Till Baernighausen, Yemane Berhane, Japhet Killewo, Ayoade M.J. Oduola, Ali Sie, Emily R Smith, Abdramane Bassiahi Soura, Raji Tajudeen, Said Vuai, Wafaie W Fawzi

## Abstract

COVID-19 vaccine hesitancy among adolescents poses a challenge to the global effort to control the pandemic. This multi-country survey aimed to assess the levels and determinants of COVID-19 vaccine hesitancy among adolescents in sub-Saharan Africa between July and December 2021. The survey was conducted using computer-assisted telephone interviewing among adolescents in five sub-Saharan African countries, Burkina Faso, Ethiopia, Ghana, Nigeria, and Tanzania. A rural area and an urban area were included in each country (except Ghana, which only had a rural area), with approximately 300 adolescents in each area and 2803 in total. Sociodemographic characteristics and perceptions and attitudes on COVID-19 vaccines were measured. Vaccine hesitancy was defined as definitely not getting vaccinated or being undecided on whether to get vaccinated if a COVID-19 vaccine were available. Log-binomial models were used to calculate the adjusted prevalence ratios (aPRs) and 95% confidence intervals (CIs) for associations between potential determinants and COVID-19 vaccine hesitancy. The percentage of COVID-19 vaccine hesitancy was 15% in rural Kersa, 24% in rural Ibadan, 31% in rural Nouna, 33% in urban Ouagadougou, 37% in urban Addis Ababa, 48% in rural Kintampo, 64% in urban Lagos, 76% in urban Dar es Salaam, and 88% in rural Dodoma. Perceived low necessity, concerns about vaccine safety, and concerns about vaccine effectiveness were the leading reasons for hesitancy. Healthcare workers, parents or family members, and schoolteachers had the greatest impacts on vaccine willingness. Perceived lack of safety (aPR: 3.61; 95% CI: 3.10, 4.22) and lack of effectiveness (aPR: 3.59; 95% CI: 3.09, 4.18) were associated with greater vaccine hesitancy. The levels of COVID-19 vaccine hesitancy among adolescents are alarmingly high across the five sub-Saharan African countries, especially in Tanzania. COVID-19 vaccination campaigns among sub-Saharan African adolescents should address their concerns and misconceptions about vaccine safety and effectiveness.

## Introduction

With unprecedented speed, multiple vaccines against SARS-CoV-2 were developed one year after the beginning of the coronavirus disease 2019 (COVID-19) pandemic. While the COVID-19 vaccination campaigns worldwide continue with the immunization of adults, COVID-19 vaccines have been or may be made available to adolescents in many settings. The United States Centers for Disease Control and Prevention currently recommends the Pfizer-BioNTech COVID-19 vaccine to children and adolescents ages 5 to 17 years old [1].

The morbidity and mortality from COVID-19 are much lower in adolescents than among adults, especially when compared to the most vulnerable group aged 65 years and older [2]. However, there is a strong rationale for providing COVID-19 vaccines to children and adolescents. Infected adolescents can still transmit the virus to other individuals [3], and some adolescents do themselves develop severe symptoms and complications from COVID-19 [4-6]. Therefore, vaccinating the adolescent population has the dual benefits of protecting adolescents against morbidity and mortality while reducing the spread of the virus by promoting herd immunity [3, 7]. In sub-Saharan Africa, adolescents aged 10 to 19 years make up 23% of the population [8]. Therefore, getting COVID-19 vaccines into the arms of adolescents is crucial for Africa to achieve the World Health Organization’s target of 70% COVID-19 vaccination coverage by mid-2022 [9]. Further, adolescents may serve as agents of advocacy that encourage their family members and friends to get vaccinated [10].

Existing evidence shows that COVID-19 vaccines are safe and effective in adolescents [11]. However, studies in high-income settings show the concerning issue of COVID-19 vaccine hesitancy among adolescents. A survey conducted by the United States Centers for Disease Control and Prevention among adolescents and their parents in April 2021 (just before the expanded availability of the Pfizer-BioNTech vaccine to adolescents) showed that only 52% of unvaccinated adolescents aged 13 to 17 years would definitely or probably receive a COVID-19 vaccine [12]. Vaccine hesitancy, defined as the reluctance to accept available vaccines [13], was considered by the World Health Organization as one of the top threats to global health even before the COVID-19 pandemic [14]. Vaccine hesitancy of adolescents poses a challenge to the global effort to control the COVID-19 pandemic. Perceptions and attitudes toward vaccines are heavily driven by cultural, social, historical, political, and individual factors that greatly vary across cultural and geographical settings [15]. As a result, COVID-19 vaccine hesitancy varies considerably across regions and countries, as shown repeatedly in previous surveys among adults [16-21]. Further, the levels and determinants of vaccine hesitancy in adults may not match those among adolescents [22].

Previous studies on COVID-19 vaccine hesitancy of adolescents were conducted in high-income countries [12, 22-25] and China [10], whereas studies in sub-Saharan African countries [19, 26-32] and other LMICs [31, 33-43] have primarily examined the adult population. To the best of our knowledge, no study has specifically examined the levels and determinants of COVID-19 vaccine hesitancy among adolescents across diverse settings in sub-Saharan Africa, a region where adolescents make up a considerable share of the population [44].

With the COVID-19 vaccination efforts around the globe moving toward children and adolescents, a better understanding of the extent and the driving factors of vaccine hesitancy in adolescents is needed to devise vaccination campaigns that increase vaccine uptake among adolescents. In this telephone survey across five sub-Saharan African countries, we aimed to assess the levels of COVID-19 vaccine hesitancy in adolescents and evaluate the potential determinants of adolescent COVID-19 vaccine hesitancy.

## Methods

### Study design and study population

This cross-country study was based on an ongoing survey that used a novel mobile platform and computer-assisted telephone interviewing to collect data from sub-Saharan African adolescents, adults, and healthcare providers. In the previous round of the survey (Round 1), six areas from three countries were included, namely Nouna and Ouagadougou in Burkina Faso, Kersa and Addis Ababa in Ethiopia, and Ibadan and Lagos in Nigeria. In each country, one rural area (Nouna, Kersa, and a rural subarea in Ibadan) and one urban area (Ouagadougou, Addis Ababa, and Lagos) were included. The details of the study design and study population in Round 1 were described previously [45]. In the currently reported new round (Round 2), a rural subarea in Kintampo (Ghana), a rural subarea in Dodoma (Tanzania), and an urban subarea in Dar es Salaam (Tanzania) were added as new participating areas. Therefore, the Round 2 survey included nine areas from five countries. Although Ibadan and Dodoma are generally considered urban, the surveys were conducted in more rural subareas of Ibadan and Dodoma. The countries and areas were selected based on existing collaborations and infrastructure available in the African Research, Implementation Science and Education (ARISE) Network [46] to represent diverse settings across sub-Saharan Africa.

The detailed study protocol of the Round 2 survey is available on the website of the Harvard University Center for African Studies (https://africa.harvard.edu/files/african-studies/files/arise_covid_survey_round_2_methods_brief_final.pdf). Briefly, during Round 2, households were selected from sampling frames of existing Health and Demographic Surveillance Systems (HDSSs) or national surveys, where possible. Within the sampling frame in each area, we interviewed approximately 300 randomly selected households with adolescents between the ages of 10 to 19 years residing in the household. The sampling frames and sampling methods for Burkina Faso, Ethiopia, and Nigeria have been described in detail previously [45]. For the new study area of Kintampo (Ghana), we used the Kintampo HDSS as the sampling frame. The Kintampo HDSS represents a catchment area of 163182 individuals and 39134 households. A total of 3589 adolescents were randomly sampled from the overall list of households, and 1074 households were called until the target sample size of 300 adolescents was reached. For the new study area of Dodoma (Tanzania), we used the Dodoma HDSS as the sampling frame; 600 households with adolescents were selected randomly from the sampling frame, and 318 eligible adolescents completed the survey. For the new study area of Dar es Salaam (Tanzania), we used the Dar es Salaam HDSS, also referred to as the Dar es Salaam Urban Cohort Study (DUCS) [47], as the sampling frame. The Dar es Salaam HDSS included 14754 households comprised of 143452 individuals, of which 30446 were adolescents. We randomly selected 2500 households, and approximately 655 households were contacted to attain the sample size of 302 adolescents.

The Round 1 survey was conducted between July and November 2020, and the Round 2 survey was conducted between July and December 2021. For the Round 2 survey, all households with adolescents aged 10 to 19 years that participated in the Round 1 survey were contacted again. A small number of participants aged 19 years during Round 1 and aged 20 years at the time of Round 2 were also eligible for inclusion to enrich the examination of the potential impacts of Round 1 predictors. For households that could not be contacted, new households were randomly identified from the sampling frame until the minimum target sample size of 300 households per area was reached. For Tanzania and Ghana, however, none of the adolescents participated in Round 1, as the two countries were added during the Round 2 survey.

Verbal parental consent and adolescent assent were obtained for adolescents younger than 18 years of age, and oral informed consent was obtained from adolescents aged 18 years and older. This study was approved by the Institutional Review Board at Harvard T.H. Chan School of Public Health and ethical review boards in each country and area, including the Nouna Health Research Center Ethical Committee and National Ethics Committee in Burkina Faso, the Institutional Ethical Review Board of Addis Continental Institute of Public Health in Ethiopia, Kintampo Health Research Centre Institutional Ethics Committee in Ghana, the University of Ibadan Research Ethics Committee and National Health Research Ethics Committee in Nigeria, and the University of Dodoma, Muhimbili University of Health and Allied Sciences, and National Institute for Medical Research in Tanzania.

### Data collection

The Round 1 survey included questions on the adolescents’ sociodemographic characteristics, knowledge and perceptions of COVID-19, and the impacts of the COVID-19 pandemic on adolescents’ education, daily activities, communication, media consumption, and various health domains, including dietary intake and mental health [48]. The Round 2 survey focused heavily on adolescents’ awareness, knowledge, perceptions, and attitudes toward COVID-19 vaccines, willingness to receive COVID-19 vaccines, potential determinants of COVID-19 vaccine hesitancy, trusted sources of information regarding COVID-19 vaccines, and the expectations on COVID-19 vaccine campaigns. The complete adolescent instrument used for the Round 2 survey is available on the website of the Harvard University Center for African Studies (https://africa.harvard.edu/files/african-studies/files/arise_covid-19_survey_round_2_adolescent_household_survey_questionnaire.pdf).

### Statistical analysis

We conducted descriptive analyses on the sociodemographic characteristics of the adolescents, willingness to receive the COVID-19 vaccine, perceptions of the vaccine, self-reported determinants of their willingness to be vaccinated, and their trusted information sources and expectations about the vaccine. For the descriptive analyses, we calculated means and standard deviations (SDs) overall and by area for continuous variables and counts and percentages for categorical variables.

The outcome for the associational analyses was COVID-19 vaccine hesitancy, defined as a response of definitely not getting the COVID-19 vaccine or maybe, unsure, or undecided on whether to get the COVID-19 vaccine if it were available now. We defined vaccine hesitancy as a dichotomous outcome. The potential determinants included age, sex, country, rural residence, currently receiving education, perceived safety of the COVID-19 vaccine among children and adolescents, perceived effectiveness of the COVID-19 vaccine, current impacts of the COVID-19 pandemic on daily activities, levels of psychological distress measured using the four-item Patient Health Questionnaire for Depression and Anxiety Scale [49], perceived risk of being exposed to COVID-19 during Round 1 (for Round 1 participants only), and knowledge score on the symptoms, transmission, and prevention of COVID-19 measured during Round 1 (for Round 1 participants only).

We used log-binomial models to calculate the prevalence ratios (PRs) and 95% confidence intervals (CIs) and modified Poisson models to achieve model convergence whenever necessary [50]. We examined the crude associations between potential determinants and vaccine hesitancy in unadjusted models. We estimated adjusted prevalence ratios (aPRs) in adjustedmodels controlled for age, sex, country, and rural residence. We conducted all analyses using SAS 9.4 (SAS Institute Inc., Cary, North Carolina) at a two-sided α level of 0.05. Missing data were handled using the complete case analysis.

## Results

### Sociodemographic characteristics

A total of 2803 adolescents were interviewed (**Table 1**). The age of the adolescents ranged from a minimum of 10 years to a maximum of 20 years. The mean age ranged from 14 years in Dodoma (SD: 2.3) and Dar es Salaam (SD: 2.6) to 17 years in Addis Ababa (SD: 2.4) and Ibadan (SD: 2.0). The sex distributions were roughly balanced between girls and boys, except for Kersa, where 31% were girls, and Addis Ababa, where 62% were girls. The percentage of adolescents with at least some secondary school education ranged from 16% in Dodoma to 96% in Ibadan. No less than 85% of the participants self-identified as students, except in Nouna and Dodoma, where 52% and 57% of the adolescents were students, respectively. In Burkina Faso, Ethiopia, and Nigeria, adolescents who also participated in the Round 1 survey varied from 9% in Lagos to 64% in Addis Ababa.

**Table 1.** Sociodemographic characteristics of adolescents in a phone-based survey in five sub-Saharan African countries, 2021^1^

|  | Burkina Faso |  | Ethiopia |  | Ghana | Nigeria |  | Tanzania |  | Total |
| --- | --- | --- | --- | --- | --- | --- | --- | --- | --- | --- |
|  | Rural | Urban | Rural | Urban | Rural | Rural | Urban | Rural | Urban |  |
|  | Nouna | Ouagadougou | Kersa | Addis Ababa | Kintampo | Ibadan | Lagos | Dodoma | Dar es Salaam |  |
| Number of adolescents, *N* | 309 | 300 | 300 | 308 | 300 | 319 | 347 | 318 | 302 | 2803 |
| Adolescents who also participated in Round 1 survey, *N* (%) | 185 (59.9) | 181 (60.3) | 109 (36.3) | 198 (64.3) | 0 (0.0) | 120 (37.6) | 31 (8.9) | 0 (0.0) | 0 (0.0) | 824 (29.4) |
| Age, years (SD) | 15.8 (2.5) | 15.8 (2.5) | 16.1 (2.8) | 17.0 (2.4) | 15.6 (2.7) | 16.9 (2.0) | 16.0 (1.9) | 14.4 (2.3) | 14.1 (2.6) | 15.7 (2.6) |
| Girls, *N* (%) | 140 (45.3) | 161 (53.7) | 92 (30.7) | 191 (62.0) | 153 (51.0) | 175 (54.9) | 206 (59.4) | 175 (55.0) | 177 (58.6) | 1470 (52.4) |
| Highest level of education,<sup>2,3</sup> *N* (%) |  |  |  |  |  |  |  |  |  |  |
| None/religious school/literacy class | 38 (12.4) | 17 (5.7) | 2 (0.7) | 1 (0.3) | 10 (3.3) | 1 (0.3) | 0 (0.0) | 34 (10.7) | 0 (0.0) | 103 (3.7) |
| Some primary school | 90 (29.4) | 58 (19.3) | 139 (46.3) | 59 (19.2) | 109 (36.3) | 3 (0.9) | 0 (0.0) | 169 (53.1) | 109 (36.5) | 736 (26.3) |
| Completed primary school | 40 (13.1) | 16 (5.3) | 30 (10.0) | 30 (9.7) | 46 (15.3) | 9 (2.8) | 17 (4.9) | 65 (20.4) | 52 (17.4) | 305 (10.9) |
| Some secondary/high school | 114 (37.3) | 186 (62.0) | 103 (34.3) | 130 (42.2) | 127 (42.3) | 216 (67.9) | 299 (86.2) | 48 (15.1) | 113 (37.8) | 1336 (47.8) |
| Completed secondary/high school | 11 (3.6) | 20 (6.7) | 10 (3.3) | 50 (16.2) | 7 (2.3) | 78 (24.5) | 27 (7.8) | 2 (0.6) | 19 (6.4) | 224 (8.0) |
| Tertiary education or higher | 13 (4.3) | 3 (1.0) | 16 (5.3) | 38 (12.3) | 1 (0.3) | 11 (3.5) | 4 (1.2) | 0 (0.0) | 6 (2.0) | 92 (3.3) |
| Occupation<sup>4</sup> N (%) |  |  |  |  |  |  |  |  |  |  |
| Unemployed | 21 (6.8) | 12 (4.0) | 2 (0.7) | 5 (1.6) | 11 (3.7) | 0 (0.0) | 0 (0.0) | 71 (22.3) | 21 (7.0) | 143 (5.1) |
| Student | 161 (52.1) | 255 (85.0) | 278 (92.7) | 290 (94.2) | 259 (86.3) | 309 (97.5) | 342 (98.6) | 181 (56.9) | 267 (88.4) | 2342 (83.6) |
| Farmer | 86 (27.8) | 0 (0.0) | 16 (5.3) | 2 (0.7) | 14 (4.7) | 0 (0.0) | 0 (0.0) | 34 (10.7) | 1 (0.3) | 153 (5.5) |
| Wage employment | 0 (0.0) | 2 (0.7) | 0 (0.0) | 5 (1.6) | 3 (1.0) | 1 (0.3) | 3 (0.9) | 0 (0.0) | 3 (1.0) | 17 (0.6) |
| Self-employed | 7 (2.3) | 9 (3.0) | 2 (0.7) | 6 (2.0) | 8 (2.7) | 6 (1.9) | 2 (0.6) | 1 (0.3) | 10 (3.3) | 51 (1.8) |
| Casual, off-farm income | 6 (1.9) | 4 (1.3) | 0 (0.0) | 0 (0.0) | 1 (0.3) | 1 (0.3) | 0 (0.0) | 31 (9.8) | 0 (0.0) | 43 (1.5) |

<sup>1</sup> Values are mean (standard deviation) for age and count (percentage) for other variables. SD, standard deviation.
<sup>2</sup> Percentages may not add up to 100% due to rounding.
<sup>3</sup> Level of education missing for 3 adolescents in Nouna, 1 adolescent in Ibadan, and 3 adolescents in Dar es Salaam.
<sup>4</sup> Occupation missing for 2 adolescents in Ibadan.

### Willingness to receive COVID-19 vaccines

Over 80% of adolescents had previously heard of COVID-19 vaccines, except in Nouna and Kersa, where 74% and 57% of the adolescents had heard of the vaccines, respectively (**Table 2**). Kersa had the highest percentage of adolescents who would definitely get vaccinated if a COVID-19 vaccine were available (86%), whereas Dodoma had the lowest percentage of willingness to definitely get vaccinated (12%). Correspondingly, Dodoma had the highest percentage of adolescents who would definitely *not* get vaccinated (79%), and Kersa had the lowest percentage (11%). Nouna had the highest percentage of adolescents who were unsure or undecided on whether they would be vaccinated if a COVID-19 vaccine were available (17%). In ascending order, the percentage of adolescents with COVID-19 vaccine hesitancy, defined as definitely not getting vaccinated or unsure/undecided, was 15% in Kersa, 24% in Ibadan, 31% in Nouna, 33% in Ouagadougou, 37% in Addis Ababa, 48% in Kintampo, 64% in Lagos, 76% in Dar es Salaam, and 88% in Dodoma. Among adolescents who were definitely or possibly willing to receive COVID-19 vaccines, keeping themselves and their families safe was the most common reason for vaccination, cited by over 92% of adolescents in all areas except in Dodoma (63%) and Dar es Salaam (89%). Parental or familial will was the second common driver for vaccination, selected by over 60% of adolescents in all areas except Addis Ababa (45%) and Dodoma (25%). Doctors’ suggestions were the third common motivation for vaccination, chosen by over 40% of adolescents in all areas except Dodoma (16%) and Dar es Salaam (35%). Among adolescents who were definitely not or possibly not willing to receive COVID-19 vaccines, the leading reasons for hesitancy were perceived low necessity (selected by 47% of adolescents overall), concerns about the safety of the vaccine (selected by 46% overall), and concerns about the effectiveness of the vaccine (selected by 12% overall). Personal liberty concerns, the influence of conspiracy theories, religious reasons, and fears of getting unlicensed, experimental, or worse-quality vaccines were not the main reasons for hesitancy, each cited by less than 3% of adolescents.

**Table 2.** Willingness to receive the COVID-19 vaccine among adolescents in a phone-based survey in five sub-Saharan African countries, 2021^1^

|  | Burkina Faso |  | Ethiopia |  | Ghana | Nigeria |  | Tanzania |  | Total |
| --- | --- | --- | --- | --- | --- | --- | --- | --- | --- | --- |
|  | Rural | Urban | Rural | Urban | Rural | Rural | Urban | Rural | Urban |  |
|  | Nouna | Ouagadougou | Kersa | Addis Ababa | Kintampo | Ibadan | Lagos | Dodoma | Dar es Salaam |  |
| Number of adolescents, *N* | 309 | 300 | 300 | 308 | 300 | 319 | 347 | 318 | 302 | 2803 |
| Having heard of COVID-19 vaccine,<sup>2</sup> *N* (%) | 225 (73.5) | 270 (90.0) | 165 (56.9) | 303 (98.4) | 261 (87.9) | 280 (90.0) | 337 (97.1) | 267 (84.0) | 295 (98.0) | 2403 (86.5) |
| Willingness to receive COVID-19 vaccine if it were available now,<sup>3,4</sup> *N* (%) |  |  |  |  |  |  |  |  |  |  |
| Would definitely not get vaccinated | 30 (13.3) | 71 (26.4) | 18 (10.9) | 89 (29.4) | 92 (35.3) | 45 (16.1) | 176 (52.2) | 210 (78.7) | 198 (67.1) | 929 (38.7) |
| Would definitely get vaccinated | 156 (69.3) | 179 (66.5) | 141 (85.5) | 191 (63.0) | 136 (52.1) | 212 (76.0) | 123 (36.5) | 32 (12.0) | 72 (24.4) | 1242 (51.7) |
| Maybe/unsure/undecided | 39 (17.3) | 19 (7.1) | 6 (3.6) | 23 (7.6) | 33 (12.6) | 22 (7.9) | 38 (11.3) | 25 (9.4) | 25 (8.5) | 230 (9.6) |
| Reasons for getting the COVID-19 vaccine,<sup>5,6</sup> *N* (%) |  |  |  |  |  |  |  |  |  |  |
| To keep self and family safe | 182 (93.3) | 195 (98.5) | 143 (97.3) | 212 (99.1) | 156 (92.3) | 225 (96.2) | 155 (96.3) | 36 (63.2) | 86 (88.7) | 1390 (94.4) |
| Parents' or family's will | 172 (88.2) | 174 (87.9) | 106 (72.1) | 96 (44.9) | 125 (74.0) | 189 (80.8) | 98 (60.9) | 14 (24.6) | 69 (71.1) | 1043 (70.9) |
| Because doctor suggested | 156 (80.0) | 192 (97.0) | 66 (44.9) | 96 (44.9) | 119 (70.4) | 172 (73.5) | 76 (47.2) | 9 (15.8) | 34 (35.1) | 920 (62.5) |
| Reasons for not getting the COVID-19 vaccine,<sup>5,7</sup> *N* (%) |  |  |  |  |  |  |  |  |  |  |
| Perceived low necessity of being vaccinated | 48 (71.6) | 52 (60.5) | 15 (62.5) | 37 (33.3) | 20 (18.0) | 22 (36.1) | 81 (38.2) | 168 (90.8) | 59 (26.8) | 502 (46.6) |
| Concerns about effectiveness of the vaccine | 9 (13.4) | 28 (32.6) | 0 (0.0) | 23 (20.7) | 7 (6.3) | 7 (11.5) | 40 (18.9) | 2 (1.1) | 8 (3.6) | 124 (11.5) |
| Concerns about safety of the vaccine | 24 (35.8) | 55 (64.0) | 9 (37.5) | 74 (66.7) | 59 (53.2) | 33 (54.1) | 146 (68.9) | 6 (3.2) | 88 (40.0) | 494 (45.9) |
| Fear of getting unlicensed/experimental/worse-quality vaccines | 2 (3.0) | 1 (1.2) | 0 (0.0) | 4 (3.6) | 3 (2.7) | 1 (1.6) | 11 (5.2) | 1 (0.5) | 2 (0.9) | 25 (2.3) |
| Religious reasons | 0 (0.0) | 0 (0.0) | 0 (0.0) | 12 (10.8) | 1 (0.9) | 0 (0.0) | 2 (0.9) | 0 (0.0) | 1 (0.5) | 16 (1.5) |
| Influence of conspiracy theories | 1 (1.5) | 1 (1.2) | 0 (0.0) | 5 (4.5) | 13 (11.7) | 2 (3.3) | 4 (1.9) | 0 (0.0) | 0 (0.0) | 26 (2.4) |
| Personal liberty concerns | 0 (0.0) | 2 (2.3) | 0 (0.0) | 2 (1.8) | 5 (4.5) | 0 (0.0) | 1 (0.5) | 2 (1.1) | 3 (1.4) | 15 (1.4) |

<sup>1</sup> Values are counts (percentages) for categorical variables. <sup>2</sup> Missing for 3 adolescents in Nouna, 10 adolescents in Kersa, 8 adolescents in Ibadan, 1 adolescent in Dar es Salaam, and 3 adolescents in Kintampo. <sup>3</sup> Percentages may not add up to 100% due to rounding. <sup>4</sup> Among adolescents who have heard of COVID-19 vaccines. Missing for 1 adolescent in Ouagadougou and 1 adolescent in Ibadan. <sup>5</sup> Counts and percentages do not add up to the total because the selection of multiple reasons was allowed. <sup>6</sup> Counts and percentages are among adolescents who would definitely get vaccinated and who were unsure/undecided. <sup>7</sup> Counts and percentages are among adolescents who would definitely not get vaccinated and who were unsure/undecided. Missing for 2 adolescents in Nouna, 4 adolescents in Ouagadougou, 1 adolescent in Addis Ababa, 6 adolescents in Ibadan, 2 adolescents in Lagos, 3 adolescents in Dar es Salaam, 50 adolescents in Dodoma, and 14 adolescents in Kintampo.

### Perceptions of COVID-19 vaccines

The percentage of adolescents who perceived COVID-19 vaccines to be very safe or somewhat safe among the general population ranged from 38% in Dodoma to 76% in Kersa and Addis Ababa (**Table 3**). Similarly, the percentage of adolescents perceiving COVID-19 vaccines as very safe or somewhat safe among children and adolescents varied from 33% in Dodoma to 77% in Kersa. In terms of the perceived effectiveness of COVID-19 vaccines among the general population, 39% of participants in Dodoma to 80% of participants in Kersa perceived COVID-19 vaccines to be very effective or somewhat effective in preventing COVID-19 infection. Overall, 46% of adolescents could not identify any possible side effects of COVID-19 vaccines, and 27% believed that there were no side effects from the vaccines.

**Table 3.** Perceptions of the COVID-19 vaccine among adolescents in a phone-based survey in five sub-Saharan African countries, 2021^1^

|  | Burkina Faso |  | Ethiopia |  | Ghana | Nigeria |  | Tanzania |  | Total |
| --- | --- | --- | --- | --- | --- | --- | --- | --- | --- | --- |
|  | Rural | Urban | Rural | Urban | Rural | Rural | Urban | Rural | Urban |  |

|  | Nouna | Ouagadougou | Kersa | Addis Ababa | Kintampo | Ibadan | Lagos | Dodoma | Dar es Salaam |  |
| --- | --- | --- | --- | --- | --- | --- | --- | --- | --- | --- |
| Number of adolescents, *N* | 309 | 300 | 300 | 308 | 300 | 319 | 347 | 318 | 302 | 2803 |
| Perceived safety of the COVID-19 vaccine in general,<sup>2,3</sup> *N* (%) |  |  |  |  |  |  |  |  |  |  |
| Very safe | 135 (43.7) | 119 (39.9) | 224 (74.7) | 50 (16.2) | 67 (22.3) | 171 (53.6) | 97 (28.0) | 46 (14.5) | 92 (30.5) | 1001 (35.7) |
| Somewhat safe | 68 (22.0) | 56 (18.8) | 4 (1.3) | 184 (59.7) | 69 (23.0) | 61 (19.1) | 90 (25.9) | 76 (23.9) | 57 (18.9) | 665 (23.7) |
| Not very safe | 50 (16.2) | 54 (18.1) | 6 (2.0) | 28 (9.1) | 42 (14.0) | 24 (7.5) | 83 (23.9) | 80 (25.2) | 28 (9.3) | 395 (14.1) |
| Not safe at all | 5 (1.6) | 20 (6.7) | 2 (0.7) | 25 (8.1) | 16 (5.3) | 12 (3.8) | 28 (8.1) | 19 (6.0) | 54 (17.9) | 181 (6.5) |
| Do not know | 51 (16.5) | 49 (16.4) | 64 (21.3) | 21 (6.8) | 106 (35.3) | 51 (16.0) | 49 (14.1) | 97 (30.5) | 71 (23.5) | 559 (20.0) |
| Perceived safety of the COVID-19 vaccine among children and adolescents,<sup>2,4</sup> *N* (%) |  |  |  |  |  |  |  |  |  |  |
| Very safe | 131 (42.4) | 91 (31.4) | 230 (76.7) | 67 (21.8) | 50 (16.7) | 151 (47.3) | 93 (26.8) | 39 (12.3) | 84 (27.8) | 936 (33.5) |
| Somewhat safe | 65 (21.0) | 62 (21.4) | 2 (0.7) | 141 (45.8) | 59 (19.7) | 67 (21.0) | 85 (24.5) | 66 (20.8) | 47 (15.6) | 594 (21.3) |
| Not very safe | 53 (17.2) | 58 (20.0) | 7 (2.3) | 42 (13.6) | 48 (16.0) | 31 (9.7) | 84 (24.2) | 84 (26.5) | 28 (9.3) | 435 (15.6) |
| Not safe at all | 7 (2.3) | 25 (8.6) | 2 (0.7) | 30 (9.7) | 27 (9.0) | 11 (3.5) | 30 (8.7) | 31 (9.8) | 66 (21.9) | 229 (8.2) |
| Do not know | 53 (17.2) | 54 (18.6) | 59 (19.7) | 28 (9.1) | 116 (38.7) | 59 (18.5) | 55 (15.9) | 97 (30.6) | 77 (25.5) | 598 (21.4) |
| Perceived effectiveness of the COVID-19 vaccine in general,<sup>2,5</sup> *N* (%) |  |  |  |  |  |  |  |  |  |  |
| Very effective | 136 (44.0) | 120 (40.8) | 220 (73.3) | 66 (21.4) | 63 (21.0) | 164 (51.4) | 88 (25.4) | 48 (15.2) | 71 (23.5) | 976 (34.9) |
| Somewhat effective | 65 (21.0) | 56 (19.1) | 20 (6.7) | 149 (48.4) | 58 (19.3) | 72 (22.6) | 81 (23.3) | 75 (23.8) | 57 (18.9) | 633 (22.7) |
| Not very effective | 42 (13.6) | 38 (12.9) | 5 (1.7) | 30 (9.7) | 31 (10.3) | 24 (7.5) | 71 (20.5) | 80 (25.4) | 31 (10.3) | 352 (12.6) |
| Not effective at all | 6 (1.9) | 20 (6.8) | 2 (0.7) | 21 (6.8) | 16 (5.3) | 5 (1.6) | 25 (7.2) | 19 (6.0) | 54 (17.9) | 168 (6.0) |
| Do not know | 60 (19.4) | 60 (20.4) | 53 (17.7) | 42 (13.6) | 132 (44.0) | 54 (16.9) | 82 (23.6) | 93 (29.5) | 89 (29.5) | 665 (23.8) |
| Perceived side effects of the COVID-19 vaccine,<sup>6,7</sup> *N* (%) |  |  |  |  |  |  |  |  |  |  |
| No side effects | 52 (16.8) | 73 (24.4) | 130 (43.3) | 105 (34.1) | 49 (16.3) | 111 (34.8) | 54 (15.6) | 99 (31.3) | 88 (29.1) | 761 (27.2) |
| Fever | 64 (20.7) | 42 (14.1) | 1 (0.3) | 69 (22.4) | 33 (11.0) | 58 (18.2) | 83 (23.9) | 3 (1.0) | 25 (8.3) | 378 (13.5) |
| Body ache including sore arm | 59 (19.1) | 19 (6.4) | 7 (2.3) | 115 (37.3) | 38 (12.7) | 74 (23.2) | 94 (27.1) | 3 (1.0) | 13 (4.3) | 422 (15.1) |
| Nausea | 54 (17.5) | 22 (7.4) | 1 (0.3) | 51 (16.6) | 10 (3.3) | 11 (3.5) | 19 (5.5) | 0 (0.0) | 6 (2.0) | 174 (6.2) |
| Tiredness/exhaustion | 62 (20.1) | 38 (12.7) | 7 (2.3) | 76 (24.7) | 24 (8.0) | 73 (22.9) | 69 (19.9) | 0 (0.0) | 10 (3.3) | 359 (12.8) |
| Do not know any | 183 (59.2) | 158 (52.8) | 158 (52.7) | 41 (13.3) | 179 (59.7) | 109 (34.2) | 124 (35.7) | 203 (64.2) | 119 (39.4) | 1274 (45.5) |

<sup>1</sup> Values are counts (percentages) for categorical variables. <sup>2</sup> Percentages may not add up to 100% due to rounding. <sup>3</sup> Missing for 2 adolescents in Ouagadougou. <sup>4</sup> Missing for 10 adolescents in Ouagadougou and 1 adolescent in Dodoma. <sup>5</sup> Missing for 6 adolescents in Ouagadougou and 3 adolescents in Dodoma. <sup>6</sup> Counts and percentages do not add up to the total because the selection of multiple reasons was allowed. <sup>7</sup> Missing for 1 adolescent in Ouagadougou and 2 adolescents in Dodoma.

### Self-reported determinants of willingness to receive vaccines

The percentage of adolescents whose willingness to receive COVID-19 vaccines would be affected by the vaccine’s country of origin ranged from 15% in Ibadan to 44% in Lagos (**Table 4**). In descending order, the overall percentages of adolescents willing to receive COVID-19 vaccines developed by specific countries were 37% for the United States, 21% for China, 15% for Europe, 13% for Russia, and 11% for India. Overall, 29% of the participants would be more willing to receive a COVID-19 vaccine developed or tested in Africa, ranging from 15% in Ibadan to 48% in Kintampo. The individuals or groups that had the greatest impacts on the adolescents’ willingness to receive COVID-19 vaccines were healthcare workers (60% overall), parents or family members (58% overall), and schoolteachers (46% overall). Religious leaders affected COVID-19 vaccine willingness for over 20% of adolescents in all areas (36% overall).

**Table 4.** Determinants of the willingness to receive the COVID-19 vaccine among adolescents in a phone-based survey in five sub-Saharan African countries, 2021^1^

|  | Burkina Faso |  | Ethiopia |  | Ghana | Nigeria |  | Tanzania |  | Total |
| --- | --- | --- | --- | --- | --- | --- | --- | --- | --- | --- |
|  | Rural | Urban | Rural | Urban | Rural | Rural | Urban | Rural | Urban |  |
|  | Nouna | Ouagadougou | Kersa | Addis Ababa | Kintampo | Ibadan | Lagos | Dodoma | Dar es Salaam |  |
| Number of adolescents, *N* | 309 | 300 | 300 | 308 | 300 | 319 | 347 | 318 | 302 | 2803 |
| Willingness affected by the vaccine's country of origin,<sup>2,3</sup> *N* (%) |  |  |  |  |  |  |  |  |  |  |
| No | 224 (72.5) | 170 (56.7) | 197 (65.7) | 217 (70.5) | 183 (61.4) | 253 (79.3) | 179 (51.7) | 210 (68.2) | 209 (69.2) | 1842 (66.0) |
| Yes | 75 (24.3) | 118 (39.3) | 58 (19.3) | 86 (27.9) | 88 (29.5) | 48 (15.1) | 152 (43.9) | 49 (15.9) | 66 (21.9) | 740 (26.5) |
| Do not know | 10 (3.2) | 12 (4.0) | 45 (15.0) | 5 (1.6) | 27 (9.1) | 18 (5.6) | 15 (4.3) | 49 (15.9) | 27 (8.9) | 208 (7.5) |
| Willing to receive COVID-19 vaccine developed by the country,<sup>4,5</sup> *N* (%) |  |  |  |  |  |  |  |  |  |  |
| United States | 36 (42.4) | 41 (31.5) | 19 (18.5) | 33 (36.3) | 53 (46.9) | 35 (53.0) | 112 (67.1) | 1 (1.1) | 14 (15.1) | 344 (36.6) |
| China | 33 (38.8) | 48 (36.9) | 27 (26.2) | 24 (26.4) | 22 (19.5) | 5 (7.6) | 28 (16.8) | 1 (1.1) | 9 (9.7) | 197 (21.0) |
| Russia | 26 (30.6) | 27 (20.8) | 6 (5.8) | 12 (13.2) | 39 (34.5) | 4 (6.1) | 10 (6.0) | 0 (0.0) | 1 (1.1) | 125 (13.3) |
| India | 15 (17.7) | 26 (20.0) | 3 (2.9) | 6 (6.6) | 29 (25.7) | 4 (6.1) | 18 (10.8) | 1 (1.1) | 2 (2.2) | 104 (11.1) |
| Europe | 18 (21.2) | 18 (13.9) | 9 (8.7) | 12 (13.2) | 29 (25.7) | 14 (21.2) | 33 (19.8) | 0 (0.0) | 3 (3.2) | 136 (14.5) |
| Do not know | 24 (28.2) | 12 (9.2) | 45 (43.7) | 19 (20.9) | 28 (24.8) | 18 (27.3) | 28 (16.8) | 56 (61.5) | 34 (36.6) | 264 (28.1) |
| Willingness of receiving a COVID-19 vaccine developed or tested in Africa,<sup>2,6</sup> *N* (%) |  |  |  |  |  |  |  |  |  |  |
| Would not affect willingness | 167 (54.2) | 123 (41.0) | 187 (62.3) | 194 (63.0) | 122 (40.7) | 227 (71.2) | 187 (53.9) | 196 (65.3) | 179 (59.3) | 1582 (56.8) |
| Would decrease willingness | 13 (4.2) | 24 (8.0) | 3 (1.0) | 48 (15.6) | 15 (5.0) | 27 (8.5) | 62 (17.9) | 6 (2.0) | 0 (0.0) | 198 (7.1) |
| Would increase willingness | 111 (36.0) | 138 (46.0) | 70 (23.3) | 58 (18.8) | 143 (47.7) | 49 (15.4) | 86 (24.8) | 55 (18.3) | 103 (34.1) | 813 (29.2) |
| Do not know | 17 (5.5) | 15 (5.0) | 40 (13.3) | 8 (2.6) | 20 (6.7) | 16 (5.0) | 12 (3.5) | 43 (14.3) | 20 (6.6) | 191 (6.9) |
| Willingness affected by individuals or groups,<sup>4</sup> *N* (%) |  |  |  |  |  |  |  |  |  |  |
| Parents or family members | 248 (80.3) | 218 (72.7) | 158 (52.7) | 110 (35.7) | 231 (77.0) | 233 (73.0) | 252 (72.6) | 42 (13.2) | 130 (43.1) | 1622 (57.9) |
| Religious leaders | 143 (46.3) | 119 (39.7) | 102 (34.0) | 90 (29.2) | 161 (53.7) | 104 (32.6) | 150 (43.2) | 68 (21.4) | 74 (24.5) | 1011 (36.1) |
| Community/tribal leaders | 113 (36.6) | 104 (34.7) | 83 (27.7) | 89 (28.9) | 141 (47.0) | 61 (19.1) | 66 (19.0) | 21 (6.6) | 19 (6.3) | 697 (24.9) |
| Political leaders | 63 (20.4) | 131 (43.7) | 75 (25.0) | 49 (15.9) | 115 (38.3) | 49 (15.4) | 56 (16.1) | 27 (8.5) | 47 (15.6) | 612 (21.8) |
| Celebrities or social media influencers | 38 (12.3) | 68 (22.7) | 21 (7.0) | 61 (19.8) | 83 (27.7) | 51 (16.0) | 64 (18.4) | 12 (3.8) | 30 (9.9) | 428 (15.3) |
| Healthcare workers | 211 (68.3) | 226 (75.3) | 158 (52.7) | 172 (55.8) | 222 (74.0) | 238 (74.6) | 244 (70.3) | 136 (42.8) | 81 (26.8) | 1688 (60.2) |
| Schoolteachers | 168 (54.4) | 167 (55.7) | 152 (50.7) | 89 (28.9) | 164 (54.7) | 163 (51.1) | 101 (29.1) | 110 (34.6) | 164 (54.3) | 1278 (45.6) |

<sup>1</sup> Values are counts (percentages) for categorical variables.
<sup>2</sup> Percentages may not add up to 100% due to rounding.
<sup>3</sup> Missing for 1 adolescent in Lagos, 10 adolescents in Dodoma, and 2 adolescents in Kintampo.
<sup>4</sup> Counts and percentages do not add up to the total because the selection of multiple reasons was allowed.
<sup>5</sup> Counts and percentages are among adolescents whose willingness to take a COVID-19 vaccine may be affected by the vaccine’s country of origin. Missing for 7 adolescents in
Dodoma and 2 adolescents in Kintampo.
<sup>6</sup> Missing for 1 adolescent in Nouna and 18 adolescents in Dodoma.

### Trusted information sources and expectations of vaccines

The most highly trusted sources of information on COVID-19 vaccines for adolescents were television, radio, or newspaper (85% overall), medical professionals (83% overall), and governmental communications (74% overall) (**S1 Table**). The adolescents’ willingness to participate in a COVID-19 vaccine trial ranged from 14% in Dodoma to 65% in Kersa. No more than 2% of the adolescents had already received a COVID-19 vaccine, except in Ibadan, where 13% had been vaccinated. Overall, 41% of the adolescents did not know when a COVID-19 vaccine would be made available to them. In particular, 70% of adolescents in Dodoma and 41% in Dar es Salaam thought that a COVID-19 vaccine would never be made available to them. Overall, 90% of adolescents believed that individuals should continue following COVID-19 preventative guidelines even after vaccines became available.

### Potential determinants of COVID-19 vaccine hesitancy

Older age was associated with a lower prevalence of COVID-19 vaccine hesitancy (aPR: 0.98; 95% CI: 0.97, 1.00) (**Table 5**). Compared to girls, boys were 8% less likely to have vaccine hesitancy (aPR: 0.92; 95% CI: 0.86, 0.98). Compared to adolescents in Burkina Faso, adolescents in Ethiopia were similarly likely to have COVID-19 vaccine hesitancy (aPR: 0.91; 95% CI: 0.75, 1.11), adolescents in Ghana were 55% more likely to have COVID-19 vaccine hesitancy (aPR: 1.55; 95% CI: 1.28, 1.86), adolescents in Nigeria were 44% more likely to have COVID-19 vaccine hesitancy (aPR: 1.44; 95% CI: 1.23, 1.68), and adolescents in Tanzania were 142% more likely to have COVID-19 vaccine hesitancy (aPR: 2.42; 95% CI: 2.11, 2.78). Rural residence was associated with an 8% lower prevalence of vaccine hesitancy (aPR: 0.92; 95% CI: 0.85, 1.00). Compared to those who perceived COVID-19 vaccines to be very safe among children and adolescents, adolescents perceiving COVID-19 vaccines to be somewhat safe, not very safe, and not safe at all had 1.8 times (aPR: 1.83; 95% CI: 1.54, 2.16), 3.5 times (aPR: 3.50; 95% CI: 3.01, 4.06), and 3.6 times (aPR: 3.61; 95% CI: 3.10, 4.22) the prevalence of COVID-19 vaccine hesitancy, respectively. Similarly, compared to adolescents who believed that COVID-19 vaccines were very effective, adolescents who perceived COVID-19 vaccines to be somewhat effective, not very effective, and not effective at all had 1.9 times (aPR: 1.94; 95% CI: 1.66, 2.28), 3.4 times (aPR: 3.37; 95% CI: 2.91, 3.90), and 3.6 times (aPR: 3.59; 95% CI: 3.09, 4.18) the prevalence of vaccine hesitancy, respectively. Experiencing any current impacts of the COVID-19 pandemic on daily activities was associated with an 11% lower prevalence of COVID-19 vaccine hesitancy (aPR: 0.89; 95% CI: 0.81, 0.97). Currently receiving education, levels of psychological distress, perceived risk of exposure to COVID-19 during Round 1, and level of COVID-19 knowledge during Round 1 were not significantly associated with COVID-19 vaccine hesitancy.

**Table 5.** Potential determinants of COVID-19 vaccine hesitancy among adolescents in a phone-based survey in five sub-Saharan African countries, 2021^1^

|  | COVID-19 vaccine hesitancy<sup>2</sup> |  |
| --- | --- | --- |
|  | Unadjusted | Adjusted |
|  | PR (95% CI) | aPR (95% CI) |
| Age, years | 0.95 (0.93, 0.96) | 0.98 (0.97, 1.00) |
| Male sex | 0.84 (0.78, 0.92) | 0.92 (0.86, 0.98) |
| Country |  |  |
| Burkina Faso | 1.00 (Ref) | 1.00 (Ref) |
| Ethiopia | 0.90 (0.75, 1.09) | 0.91 (0.75, 1.11) |
| Ghana | 1.49 (1.24, 1.78) | 1.55 (1.28, 1.86) |
| Nigeria | 1.42 (1.21, 1.65) | 1.44 (1.23, 1.68) |
| Tanzania | 2.53 (2.21, 2.89) | 2.42 (2.11, 2.78) |
| Rural residence | 0.82 (0.75, 0.89) | 0.92 (0.85, 1.00) |
| Not currently receiving education | 0.81 (0.70, 0.93) | 1.04 (0.92, 1.19) |
| Perceived safety of the COVID-19 vaccine among children and adolescents |  |  |
| Very safe | 1.00 (Ref) | 1.00 (Ref) |
| Somewhat safe | 1.93 (1.61, 2.30) | 1.83 (1.54, 2.16) |
| Not very safe | 3.89 (3.34, 4.53) | 3.50 (3.01, 4.06) |
| Not safe at all | 4.64 (4.01, 5.38) | 3.61 (3.10, 4.22) |
| Do not know | 3.33 (2.85, 3.89) | 2.82 (2.42, 3.30) |
| Perceived effectiveness of the COVID-19 vaccine |  |  |
| Very effective | 1.00 (Ref) | 1.00 (Ref) |
| Somewhat effective | 2.09 (1.77, 2.46) | 1.94 (1.66, 2.28) |
| Not very effective | 3.93 (3.39, 4.55) | 3.37 (2.91, 3.90) |
| Not effective at all | 4.69 (4.08, 5.40) | 3.59 (3.09, 4.18) |
| Do not know | 3.33 (2.87, 3.87) | 2.82 (2.43, 3.28) |
| Current impacts of the COVID-19 pandemic on daily activities |  |  |
| No impacts | 1.00 (Ref) | 1.00 (Ref) |
| Some impacts | 1.17 (1.07, 1.28) | 0.89 (0.81, 0.97) |
| Psychological distress<sup>3</sup> |  |  |
| No psychological distress | 1.00 (Ref) | 1.00 (Ref) |
| Mild psychological distress | 0.96 (0.85, 1.09) | 1.03 (0.93, 1.14) |
| Moderate psychological distress | 0.93 (0.74, 1.16) | 0.99 (0.82, 1.20) |
| Severe psychological distress | 0.80 (0.45, 1.42) | 0.89 (0.55, 1.43) |
| Perceived risk of exposure to COVID-19 during Round 1<sup>4</sup> |  |  |
| No risk | 1.00 (Ref) | 1.00 (Ref) |
| Low risk | 0.92 (0.68, 1.25) | 0.87 (0.64, 1.18) |
| High risk | 0.78 (0.55, 1.11) | 0.84 (0.58, 1.21) |
| Very high risk | 0.72 (0.41, 1.28) | 0.70 (0.40, 1.24) |
| High knowledge score of COVID-19 during Round 1<sup>5</sup> | 1.01 (0.80, 1.27) | 0.90 (0.72, 1.14) |

## Discussion

This multi-country survey finds high COVID-19 vaccine hesitancy among adolescents from the nine areas across five countries in sub-Saharan Africa. The level of hesitancy is exceptionally high in Tanzania. Individual characteristics that potentially increase vaccine hesitancy are female sex, perceived lack of safety, and perceived lack of effectiveness of COVID-19 vaccines.

COVID-19 vaccine perceptions and hesitancy are influenced by social and cultural factors that vary considerably across settings [15-21]. Of note, a large sample of 13,426 participants from the general population in 19 countries showed high heterogeneity across countries in the determinants, and even the directions of associations, of vaccine acceptance [16]. This survey is, to our knowledge, the first study that examined the levels and determinants of COVID-19 vaccine hesitancy among adolescents across various settings in sub-Saharan Africa. While the proportion of adolescents with COVID-19 vaccine hesitancy is high across areas, considerable variation is also noted, ranging from 15% in Kersa to 88% in Dodoma. Previous studies in high-or upper-middle-income settings report similarly heterogeneous levels of hesitancy. A study in adolescents aged 12 to 15 years in Arkansas, United States, reported that 58% were hesitant about getting a COVID-19 vaccine [22]. A study among Canadian adolescents aged 14 to 17 years reported that 35% would not get a COVID-19 vaccine or were unsure [24]. Based on a survey conducted by the United States Centers for Disease Control and Prevention in April 2021, only 52% of unvaccinated adolescents aged 13 to 17 years would definitely or probably receive COVID-19 vaccination [12]. A study among Swedish adolescents showed that nearly one in three adolescents had not decided if they wanted to get a COVID-19 vaccine and that only 54% were willing to be vaccinated [25]. In a survey among Chinese adolescents, 76% of the participants would accept future COVID-19 vaccination [10]. In our survey, the exceptionally high level of vaccine hesitancy in Dodoma and Dar es Salaam is perhaps attributable to the misconceptions regarding COVID-19 and its vaccination, partially exacerbated by misinformation among communities in 2020 and early 2021 [51, 52].

We find that healthcare providers are adolescents’ most trusted individuals for information on COVID-19 vaccines and may have the greatest impact on the adolescents’ willingness to vaccinate. Even in Tanzania, where vaccine hesitancy is high, over 60% of adolescents trust vaccine information from healthcare providers. Previous global surveys among the general population also consistently showed that health workers are the most trusted sources of guidance about COVID-19 vaccines [31]. A survey among Turkish adults reported that adults with higher trust in health professionals had more favorable attitudes towards COVID-19 vaccines [43]. Our survey also suggests that parents’, friends’, and schoolteachers’ attitudes toward COVID-19 vaccination may influence adolescents’ willingness to get vaccinated. Similarly, a diverse sample of adolescents in the United States showed that parent and peer norms were distinct predictors of adolescent willingness to receive vaccines [23]. Therefore, healthcare workers, parents, schoolteachers, and peers should be leveraged as advocates of COVID-19 vaccines. Efforts are needed to increase vaccine acceptance among healthcare workers and adult community members in general.

Our survey finds that, in all study areas, over 40% of adolescents considered religious bodies or leaders a trusted source of information on COVID-19 vaccines and that religious leaders affected COVID-19 vaccine willingness for over 20% of adolescents. There is a mixed body of evidence regarding religious influence on COVID-19 vaccine perceptions. A survey among Turkish adults found no significant associations between religious attitudes and COVID-19 vaccine perceptions [34]. However, a survey in Pakistan showed a strong association between religious inhibitions (the belief that trust in God was sufficient to protect one from infection) and vaccine hesitancy in adults [36]. In our survey, for 15% of adolescents overall, vaccine willingness can be affected by celebrities and social media influencers. The uncritical consumption of social media may promote vaccine hesitancy [53]. In particular, emerging social media platforms may be used to spread anti-vaccination content that jeopardizes vaccine uptake, particularly for young people who are more likely to use such platforms [54]. Social media may also spread misinformation or conspiracy theories about COVID-19. In a survey among adults in Jordan, participants who believed in COVID-19 conspiracy theories were less likely to accept COVID-19 vaccines [37]. Future efforts are needed to leverage religious leaders and social media platforms as potential mechanisms for promoting adolescents’ uptake of COVID-19 vaccines.

We report that the vaccine willingness of over 15% of adolescents in all areas is affected by the vaccine’s country of origin, with COVID-19 vaccines developed in the United States the most accepted, followed by those developed in China; vaccines developed in Russia and India appear the least accepted. Among adults in Kazakhstan [35] and Zimbabwe [27], vaccine hesitancy is likewise influenced by the vaccines’ country of origin or manufacturer. In Kazakhstan, for instance, 78% of the respondents had the highest confidence in German-produced vaccines and the lowest confidence in vaccines produced in India [35]. A global survey of 19,714 adults from 17 countries showed that 52% of the respondents would only accept COVID-19 vaccines from a specific country of origin [20]. In our survey, over 15% of the adolescents in all areas are more willing to receive COVID-19 vaccines developed or tested in Africa, highlighting the importance of technology transfer for COVID-19 vaccines to increase sub-Saharan African countries’ local capacities.

Concerns about the safety and effectiveness of COVID-19 vaccines are among the top reasons for hesitancy in this survey. Accordingly, perceived lack of safety and perceived lack of effectiveness strongly predict greater vaccine hesitancy. These findings align with previous studies among the adult population in both high-income and low- and middle-income settings [20, 31, 55]. A multi-country study assessed COVID-19 vaccine acceptance across 15 general population samples, including 44,260 individuals from the United States, Russia, and 10 LMICs in Asia, Africa, and South America [31]. The survey found that vaccine acceptance in LMICs is primarily driven by the desire for personal protection, and the most commonly cited reason for hesitancy was the concern about the side effects of the vaccine [31]. Among Chinese adolescents, those who thought that COVID-19 vaccines could protect them from COVID-19 infection and those who believed that vaccines were safe were more likely to be willing to receive COVID-19 vaccines [10]. Therefore, public health campaigns for adolescents and their guardians to boost COVID-19 vaccine uptake should emphasize that the vaccines are safe for adolescents and confer protection against COVID-19 infections [11].

We find that adolescent boys have less COVID-19 vaccine hesitancy than adolescent girls, consistent with existing evidence. Among Turkish adults, males had a higher probability of having positive attitudes toward COVID-19 vaccines [34]. Similarly, in a mixed-methods study in Sweden, girls had higher anxiety levels about the vaccine than boys. The specific mechanisms for the greater vaccine hesitancy among adolescent girls are unclear but may be related to the lower tendency of risk-taking among adolescent girls than among boys (i.e., considering receiving COVID-19 vaccines as a risk-taking behavior) [56]. These findings highlight the need to address COVID-19 vaccine hesitancy, especially among adolescent girls. A study among 472,521 adults in Latin America and the Caribbean found that having depressive symptoms was associated with greater fear of the adverse effects of the COVID-19 vaccines [40], indicating that vaccine hesitancy for some individuals may be related to underlying mental health issues that must be addressed accordingly. We do not find associations between psychological distress and COVID-19 vaccine hesitancy. The impacts of the COVID-19 pandemic on the long-term mental well-being of sub-Saharan African adolescents need to be investigated further.

The strengths of this study are the inclusion of multiple countries and cities in sub-Saharan Africa and the measurement of a wide array of potential determinants of COVID-19 vaccineB hesitancy. A potential limitation of the study was that the study areas were selected based on existing connections and infrastructure, and the adolescents in each area were not selected probabilistically. Therefore, the results from this study are not expected to be representative of all adolescents in the country. Nevertheless, we increased the representativeness of the study population by including countries and areas geographically spread across sub-Saharan Africa. Therefore, the results provide important insights into the levels of determinants of COVID-19 vaccine hesitancy in sub-Saharan Africa. Another limitation is that, due to the nature of phone-based interviews, the adolescents included in most areas were those who resided in households with access to a mobile phone. Consequently, the sample might underrepresent adolescents from under-resourced households, which may affect generalizability. The presence of unreachable phone numbers and failures to pick up phone calls may also have impacts on the generalizability of the findings.

In conclusion, we show that COVID-19 vaccine hesitancy among adolescents is high across the five sub-Saharan African countries, especially in Tanzania. COVID-19 vaccination campaigns among sub-Saharan African adolescents must address adolescents’ concerns and misconceptions about COVID-19 vaccines, especially regarding vaccination safety and effectiveness. It is unfortunate that, at the time of this survey, a considerable proportion of adolescents in sub-Saharan Africa did not yet know when COVID-19 vaccines would become available to them. It is crucial to ensure that vaccines are accessible should adolescents desire to be vaccinated. It rests upon the global medical community to get the shots into the arms of the often-neglected population of sub-Saharan African adolescents.

## Data Availability

Individual participant data cannot be shared because of the requirements for ethical approvals and data transfer agreements in this multi-country study. The de-identified dataset supporting this research may be made available following a submitted request to and the completion of ethical approval and data transfer agreement from the ethical review authority in each country and site.
The complete adolescent instrument used for the Round 2 survey is available on the website of the Harvard University Center for African Studies (https://africa.harvard.edu/files/african-studies/files/arise_covid-19_survey_round_2_adolescent_household_survey_questionnaire.pdf).

## Acknowledgments

This work was supported by institutional support from Harvard T.H. Chan School of Public Health, Boston, MA; Harvard University Center for African Studies, Boston, MA; Heidelberg Institute of Global Health, Germany, and the George Washington University Milken Institute School of Public Health, Washington, DC.

We thank the study participants and data collectors for making this study possible. The survey team in Ghana is grateful for support from the Kintampo Health Research Centre of Ghana Health Service and the community leadership of Kintampo North Municipality and Kintampo South District.

## porting information

**S1 Table.Trusted information sources and expectations about the COVID-19 vaccine among adolescents hone-based survey in five sub-Saharan African countries, 2021**

## References

1. COVID-19 Vaccines for Children and Teens: National Center for Immunization and Respiratory Diseases (NCIRD), Division of Viral Diseases; 2022 [Available from: https://www.cdc.gov/coronavirus/2019-ncov/vaccines/recommendations/children-teens.html.

2. Viner RM, Mytton OT, Bonell C, Melendez-Torres GJ, Ward J, Hudson L, et al. Susceptibility to SARS-CoV-2 Infection Among Children and Adolescents Compared With Adults: A Systematic Review and Meta-analysis. JAMA pediatrics. 2021;175(2):143–56.

3. Zimet GD, Silverman RD, Fortenberry JD. Coronavirus Disease 2019 and Vaccination of Children and Adolescents: Prospects and Challenges. The Journal of pediatrics. 2021;231:254–8.

4. Ng KF, Kothari T, Bandi S, Bird PW, Goyal K, Zoha M, et al. COVID-19 multisystem inflammatory syndrome in three teenagers with confirmed SARS-CoV-2 infection. Journal of medical virology. 2020.

5. Idele P, Anthony D, Mofenson LM, Requejo J, You D, Luo C, et al. The evolving epidemiologic and clinical picture of SARS-CoV-2 and COVID-19 disease in children and young people. 2020.

6. Feldstein LR, Rose EB, Horwitz SM, Collins JP, Newhams MM, Son MBF, et al. Multisystem inflammatory syndrome in US children and adolescents. New England Journal of Medicine. 2020;383(4):334–46.

7. Klass P, Ratner AJ. Vaccinating Children against Covid-19 — The Lessons of Measles. The New England journal of medicine. 2021;384(7):589–91.

8. United Nations Department of Economic and Social Affairs Population Division. World Population Prospects 2019, Online Edition. Rev. 1. 2019.

9. World Health Organization. Achieving 70% COVID-19 Immunization Coverage by Mid-2022 2021 [Available from: https://www.who.int/news/item/23-12-2021-achieving-70-covid-19-immunization-coverage-by-mid-2022.

10. Cai H, Bai W, Liu S, Liu H, Chen X, Qi H, et al. Attitudes Toward COVID-19 Vaccines in Chinese Adolescents. Frontiers in medicine. 2021;8:691079-.

11. Lv M, Luo X, Shen Q, Lei R, Liu X, Liu E, et al. Safety, Immunogenicity, and Efficacy of COVID-19 Vaccines in Children and Adolescents: A Systematic Review. Vaccines (Basel). 2021;9(10):1102.

12. Scherer AM, Gedlinske AM, Parker AM, Gidengil CA, Askelson NM, Petersen CA, et al. Acceptability of Adolescent COVID-19 Vaccination Among Adolescents and Parents of Adolescents — United States, April 15–23, 2021. Morbidity and Mortality Weekly Report. 2021;70(28):997–1003.

13. MacDonald NE. Vaccine hesitancy: Definition, scope and determinants. Vaccine. 2015;33(34):4161–4.

14. Qayum I. Top ten global health threats for 2019: the WHO list. Journal of Rehman Medical Institute. 2019;5(2):01–2.

15. Ekwebelem OC, Yunusa I, Onyeaka H, Ekwebelem NC, Nnorom-Dike O. COVID-19 vaccine rollout: will it affect the rates of vaccine hesitancy in Africa? Public health (London). 2021;197:e18–e9.

16. Lazarus JV, Wyka K, Rauh L, Rabin K, Ratzan S, Gostin LO, et al. Hesitant or Not? The Association of Age, Gender, and Education with Potential Acceptance of a COVID-19 Vaccine: A Country-level Analysis. Journal of health communication. 2020;25(10):799–807.

17. Wake AD. The Willingness to Receive COVID-19 Vaccine and Its Associated Factors: “Vaccination Refusal Could Prolong the War of This Pandemic” – A Systematic Review. Risk management and healthcare policy. 2021;14:2609–23.

18. Sallam M. COVID-19 Vaccine Hesitancy Worldwide: A Concise Systematic Review of Vaccine Acceptance Rates. Vaccines (Basel). 2021;9(2):160.

19. Wake AD. The Acceptance Rate Toward COVID-19 Vaccine in Africa: A Systematic Review and Meta-analysis. Global pediatric health. 2021;8:2333794–X211048738.

20. Wong LP, Alias H, Danaee M, Ahmed J, Lachyan A, Cai CZ, et al. COVID-19 vaccination intention and vaccine characteristics influencing vaccination acceptance: a global survey of 17 countries. Infectious diseases of poverty. 2021;10(1):1–122.

21. Lazarus JV, Ratzan SC, Palayew A, Gostin LO, Larson HJ, Rabin K, et al. A global survey of potential acceptance of a COVID-19 vaccine. Nature medicine. 2021;27(2):225–8.

22. Willis DE, Presley J, Williams M, Zaller N, McElfish PA. COVID-19 vaccine hesitancy among youth. Human vaccines & immunotherapeutics.ahead-of-print(ahead-of-print):1-3.

23. Rogers AA, Cook RE, Button JA. Parent and Peer Norms are Unique Correlates of COVID-19 Vaccine Intentions in a Diverse Sample of U.S. Adolescents. Journal of adolescent health. 2021.

24. Afifi TO, Salmon S, Taillieu T, Stewart-Tufescu A, Fortier J, Driedger SM. Older adolescents and young adults willingness to receive the COVID-19 vaccine: Implications for informing public health strategies. Vaccine. 2021;39(26):3473–9.

25. Nilsson S, Mattson J, Berghammer M, Brorsson AL, Forsner M, Jenholt Nolbris M, et al. To be or not to be vaccinated against COVID-19 – The adolescents’ perspective – A mixed-methods study in Sweden. Vaccine: X. 2021;9:100117-.

26. Carcelen AC, Prosperi C, Mutembo S, Chongwe G, Mwansa FD, Ndubani P, et al. COVID-19 vaccine hesitancy in Zambia: a glimpse at the possible challenges ahead for COVID-19 vaccination rollout in sub-Saharan Africa. Human vaccines & immunotherapeutics.ahead-of-print(ahead-of-print):1-6.

27. McAbee L, Tapera O, Kanyangarara M. Factors Associated with COVID-19 Vaccine Intentions in Eastern Zimbabwe: A Cross-Sectional Study. Vaccines (Basel). 2021;9(10):1109.

28. Echoru I, Ajambo PD, Keirania E, Bukenya EEM. Sociodemographic factors associated with acceptance of COVID-19 vaccine and clinical trials in Uganda: a cross-sectional study in western Uganda. BMC public health. 2021;21(1):1106-.

29. Dinga JN, Sinda LK, Titanji VPK. Assessment of Vaccine Hesitancy to a COVID-19 Vaccine in Cameroonian Adults and Its Global Implication. Vaccines (Basel). 2021;9(2):175.

30. Acheampong T, Akorsikumah EA, Osae-Kwapong J, Khalid M, Appiah A, Amuasi JH. Examining Vaccine Hesitancy in Sub-Saharan Africa: A Survey of the Knowledge and Attitudes among Adults to Receive COVID-19 Vaccines in Ghana. Vaccines (Basel). 2021;9(8):814.

31. Solís Arce JS, Warren SS, Meriggi NF, Scacco A, McMurry N, Voors M, et al. COVID-19 vaccine acceptance and hesitancy in low- and middle-income countries. Nature medicine. 2021;27(8):1385–94.

32. Adebisi YA, Alaran AJ, Bolarinwa OA, Akande-Sholabi W, Lucero-Prisno III DE. When it is available, will we take it? Social media users’ perception of hypothetical COVID-19 vaccine in Nigeria. The Pan African medical journal. 2021;38:230-.

33. Harapan H, Wagner AL, Yufika A, Winardi W, Anwar S, Gan AK, et al. Acceptance of a COVID-19 Vaccine in Southeast Asia: A Cross-Sectional Study in Indonesia. Frontiers in public health. 2020;8:381-.

34. Kilic M, Ustundag Ocal N, Uslukilic G. The relationship of Covid-19 vaccine attitude with life satisfaction, religious attitude and Covid-19 avoidance in Turkey. Human vaccines & immunotherapeutics. 2021;17(10):3384–93.

35. Issanov A, Akhmetzhanova Z, Riethmacher D, Aljofan M. Knowledge, attitude, and practice toward COVID-19 vaccination in Kazakhstan: a cross-sectional study. Human vaccines & immunotherapeutics. 2021;17(10):3394–400.

36. Ahmed TF, Ahmed A, Ahmed S, Ahmed HU. Understanding COVID-19 vaccine acceptance in Pakistan: an echo of previous immunizations or prospect of change? Expert review of vaccines. 2021;20(9):1185–93.

37. El-Elimat T, AbuAlSamen MM, Almomani BA, Al-Sawalha NA, Alali FQ. Acceptance and attitudes toward COVID-19 vaccines: A cross-sectional study from Jordan. PloS one. 2021;16(4):e0250555–e.

38. Bagateli LE, Saeki EY, Fadda M, Agostoni C, Marchisio P, Milani GP. COVID-19 Vaccine Hesitancy among Parents of Children and Adolescents Living in Brazil. Vaccines (Basel). 2021;9(10):1115.

39. Abdulah DM. Prevalence and correlates of COVID-19 vaccine hesitancy in the general public in Iraqi Kurdistan: A cross-sectional study. Journal of medical virology. 2021;93(12):6722–31.

40. Urrunaga-Pastor D, Bendezu-Quispe G, Herrera-Añazco P, Uyen-Cateriano A, Toro-Huamanchumo CJ, Rodriguez-Morales AJ, et al. Cross-sectional analysis of COVID-19 vaccine intention, perceptions and hesitancy across Latin America and the Caribbean. Travel medicine and infectious disease. 2021;41:102059-.

41. Elhadi M, Alsoufi A, Alhadi A, Hmeida A, Alshareea E, Dokali M, et al. Knowledge, attitude, and acceptance of healthcare workers and the public regarding the COVID-19 vaccine: a cross-sectional study. BMC public health. 2021;21(1):955-.

42. Kumari A, Ranjan P, Chopra S, Kaur D, Kaur T, Kalanidhi KB, et al. What Indians Think of the COVID-19 vaccine: A qualitative study comprising focus group discussions and thematic analysis. Diabetes & metabolic syndrome clinical research & reviews. 2021;15(3):679–82.

43. Karabela şN, Coşkun F, Hoşgör H. Investigation of the relationships between perceived causes of COVID-19, attitudes towards vaccine and level of trust in information sources from the perspective of Infodemic: the case of Turkey. BMC public health. 2021;21(1):1195-.

44. Hervish A, Clifton D. adolescents and Young people in Sub-Saharan Africa: opportunities and challenges: UNFPA; 2012.

45. Hemler EC, Korte ML, Lankoande B, Millogo O, Assefa N, Chukwu A, et al. Design and Field Methods of the ARISE Network COVID-19 Rapid Monitoring Survey. The American journal of tropical medicine and hygiene. 2021;105(2):310.

46. Darling AM, Assefa N, Bärnighausen T, Berhane Y, Canavan CR, Guwatudde D, et al. Design and field methods of the ARISE Network Adolescent Health Study. Tropical Medicine & International Health. 2020;25(1):5–14.

47. Leyna GH, Berkman LF, Njelekela MA, Kazonda P, Irema K, Fawzi W, et al. Profile: the Dar Es Salaam health and demographic surveillance system (Dar es Salaam HDSS). International Journal of Epidemiology. 2017;46(3):801–8.

48. Wang D, Chukwu A, Millogo O, Assefa N, James C, Young T, et al. The COVID-19 Pandemic and Adolescents’ Experience in Sub-Saharan Africa: A Cross-Country Study Using a Telephone Survey. The American journal of tropical medicine and hygiene. 2021;105(2):331–41.

49. Kroenke K, Spitzer RL, Williams JB, Löwe B. An ultra-brief screening scale for anxiety and depression: the PHQ–4. Psychosomatics. 2009;50(6):613–21.

50. Zou G. A modified poisson regression approach to prospective studies with binary data. American journal of epidemiology. 2004;159(7):702–6.

51. Makoni M. Tanzania refuses COVID-19 vaccines. The Lancet (British edition). 2021;397(10274):566-.

52. Buguzi S. Covid-19: Counting the cost of denial in Tanzania. BMJ (Online). 2021;373:n1052-n.

53. Allington D, McAndrew S, Moxham-Hall VL, Duffy B. Media usage predicts intention to be vaccinated against SARS-CoV-2 in the US and the UK. Vaccine. 2021;39(18):2595–603.

54. Basch CH, Meleo-Erwin Z, Fera J, Jaime C, Basch CE. A global pandemic in the time of viral memes: COVID-19 vaccine misinformation and disinformation on TikTok. Human vaccines & immunotherapeutics. 2021;17(8):2373–7.

55. Bono SA, Faria de Moura Villela E, Siau CS, Chen WS, Pengpid S, Hasan MT, et al. Factors Affecting COVID-19 Vaccine Acceptance: An International Survey among Low-and Middle-Income Countries. Vaccines (Basel). 2021;9(5):515.

56. Reniers RLEP, Murphy L, Lin A, Bartolome SP, Wood SJ. Risk Perception and Risk-Taking Behaviour during Adolescence: The Influence of Personality and Gender. PloS one. 2016;11(4):e0153842–e.

